# Surgically Modifiable Insertion Geometry Drives Thrombogenic Flow in the Modified Blalock-Taussig-Thomas Shunt

**DOI:** 10.64898/2026.09.11.26362753

**Authors:** Yi Qiao, Ethan Penn, Jacob Miller, Scott Bugenhagen, Ram Rohatgi, Kelsey Mercer, Blaire Kulp, Pirooz Eghtesady, Guy M. Genin, David Bark, Edon J Rabinowitz

## Abstract

**Background:** The modified Blalock-Taussig-Thomas shunt (mBTTS) sustains pulmonary blood flow in infants with cyanotic or single-ventricle heart disease, but shunt thrombosis occurs in 8-12% of cases and carries substantial mortality. Systemic anticoagulation has limited efficacy.

**Objectives:** This study tested whether surgically modifiable shunt geometry might form a second, patient-specific approach to maintaining shunt patency, via management of thrombogenic hemodynamics.

**Methods:** Three-dimensional mBTTS anatomies were reconstructed from computed tomography in 10 infants (4 thrombosed, 6 patent). Pulsatile inlet waveforms derived from Doppler ultrasound were coupled with three-element Windkessel outlet models individually calibrated to catheter-derived pressures. Transient computational fluid dynamics simulations predicted wall shear rate (WSR) and elongational strain rate (ESR). Associations between surgically relevant geometric parameters and hemodynamic metrics were evaluated.

**Results:** Simulated pressures closely matched clinical measurements at all four outlets in all 10 models. Despite substantial anatomic variability, peak WSR and ESR consistently localized to the shunt-subclavian junction in 8 of 10 patients, including all 4 thrombosed shunts. Greater deviation of shunt insertion angle from perpendicular was associated with higher systolic and cycle-averaged normalized WSR (ρ = 0.94, p < 0.001; ρ = 0.88, p = 0.002) and ESR (ρ = 0.82, p = 0.007; ρ = 0.70, p = 0.03). Larger shunt diameter was associated with lower WSR and ESR. Thrombosed shunts demonstrated higher shear-related metrics than patent shunts, particularly during systole.

**Conclusions:** In patient-specific mBTTS anatomies, insertion angle and shunt diameter are determinants of local abnormal flow, concentrated at the shunt-subclavian junction. Patient-specific hemodynamic assessment may inform shunt construction and interstage risk stratification.

**CLINICAL PERSEPECTIVE:** *What is new?:*

- In patient-specific computational models of the modified Blalock-Taussig-Thomas shunt, thrombogenic blood flow patterns localize to the shunt-subclavian junction across anatomically diverse infants, identifying this as the key region of biomechanical vulnerability for deadly thrombus initiation.
- Surgically modifiable parameters, including insertion angle and shunt diameter, were identified as key determinants of local shear stress exposure.
- Thrombosed shunts exhibit higher cycle-averaged shear exposure than patent shunts, suggesting that cumulative hemodynamic burden may discriminate thrombosis risk.

*What are the clinical implications?:*

- Infants with mBTTS are among the most heavily anticoagulated patients in pediatric cardiology, but anticoagulants do not meaningfully improve outcomes. This study shows that this is because systemic anticoagulation cannot modify the local mechanical environment that initiates clot formation at the shunt interface.
- Preoperative patient-specific computational modeling may help optimize shunt configuration to reduce thrombogenic risk.
- Geometry-informed surgical planning could complement pharmacologic strategies and advance personalized care in congenital heart palliation.

## INTRODUCTION

The modified Blalock-Taussig-Thomas shunt (mBTTS) remains an essential life-saving palliative conduit for infants with single-ventricle or cyanotic congenital heart defects, providing the pulmonary blood flow necessary to bridge these patients to subsequent staged repair.^1,2^ However, shunt thrombosis continues to occur in 8-12% of cases and carries devastating consequences: five-year survival among infants who experience shunt complications falls to 33-40%, compared with 77% in those who do not.^1–8^ These outcomes persist despite aggressive systemic anticoagulation protocols that place mBTTS infants amongst the most heavily anticoagulated patients in all of pediatric cardiology.^3^ Pharmacological management has been pushed to the safety ceiling without resolving the problem, pointing to a fundamental limitation of the current treatment strategy: systemic anticoagulation acts on circulating coagulation factors and platelet reactivity but does not modify the local mechanical environment at the shunt interface, where thrombosis originates.^9–12^

A biomechanical basis for targeting this local environment is well established. Abrupt geometric transitions at vascular anastomoses generate regions of elevated wall shear and elongational flow, conditions known to unfold von Willebrand factor, promote platelet activation and aggregation, and initiate thrombus formation through shear-mediated mechanotransduction.^9,10,13–23^ The mBTTS, by design, introduces a transition of this character between the systemic and pulmonary circulations. Idealized computational modeling, including our own prior work, has suggested that shunt insertion geometry and graft dimensions impact local thrombogenic potential.^24,25^ However, idealized geometries cannot capture the anatomic asymmetry and interpatient geometric and flow variability that characterize real surgical configurations, leaving open whether these geometric effects persist and remain actionable *in vivo*.

Whether surgically modifiable geometric features continue to shape thrombogenic hemodynamics in anatomically realistic mBTTS configurations has not been established. Here, we address this using patient-specific computational fluid dynamics (CFD) in 10 mBTTS anatomies reconstructed from clinical computed tomography imaging, including both thrombosed and patent shunts, with pulsatile boundary conditions calibrated to catheter-derived pressures. By relating local hemodynamic exposure to measured geometric parameters across this cohort, we establish a translational link between surgical geometry, local flow, and thrombosis outcomes that supports geometry-informed surgical planning as a complement to pharmacologic prevention.

## METHODS

### Clinical Dataset and Model Generation

Building on our previous identification of optimized mBTTS design that reduce thrombosis risk^24^, we performed CFD simulations on patient-specific models to quantify key hemodynamic metrics and evaluate whether the previously identified mBTTS design principles hold true in real surgical anatomies. This study was approved by the Washington University IRB (ID# 202303162).

Three-dimensional (3D) patient-specific mBTTS anatomies were reconstructed from CT imaging using SimVascular (Palo Alto, CA), which incorporates the Vascular Modeling Toolkit (VMTK) for modeling and meshing (**Figure 1A**^25^).^26^ The constructed geometries are depicted in **Figure 2**. The clinical dataset comprised 10 patients treated at St. Louis Children’s Hospital (**Table 1**) and included hemodynamic data from cardiac catheterization and Doppler ultrasound studies. For each patient, time-resolved velocity waveforms were recorded at the innominate artery, shunt, and pulmonary arteries. At least three cardiac cycles were ensemble-averaged at each site to minimize measurement variability. These clinical data were used both to prescribe boundary conditions and to iteratively calibrate Windkessel parameters against catheter-measured pressures.^26–28^

**Figure 1:**
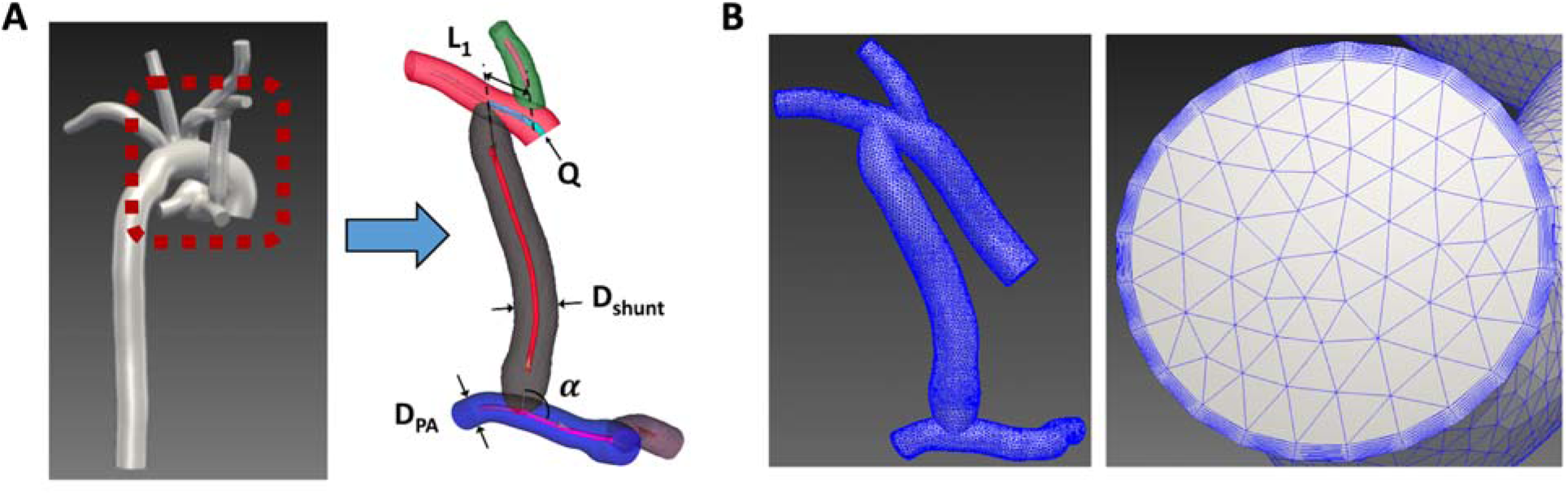
Development and Characterization of the Patient-Specific mBTTS Computational Model. **A,** Patient-specific mBTTS geometry reconstructed from CT imaging. **B,** computational mesh of the full model with detailed boundary layer refinement.

**Figure 2:**
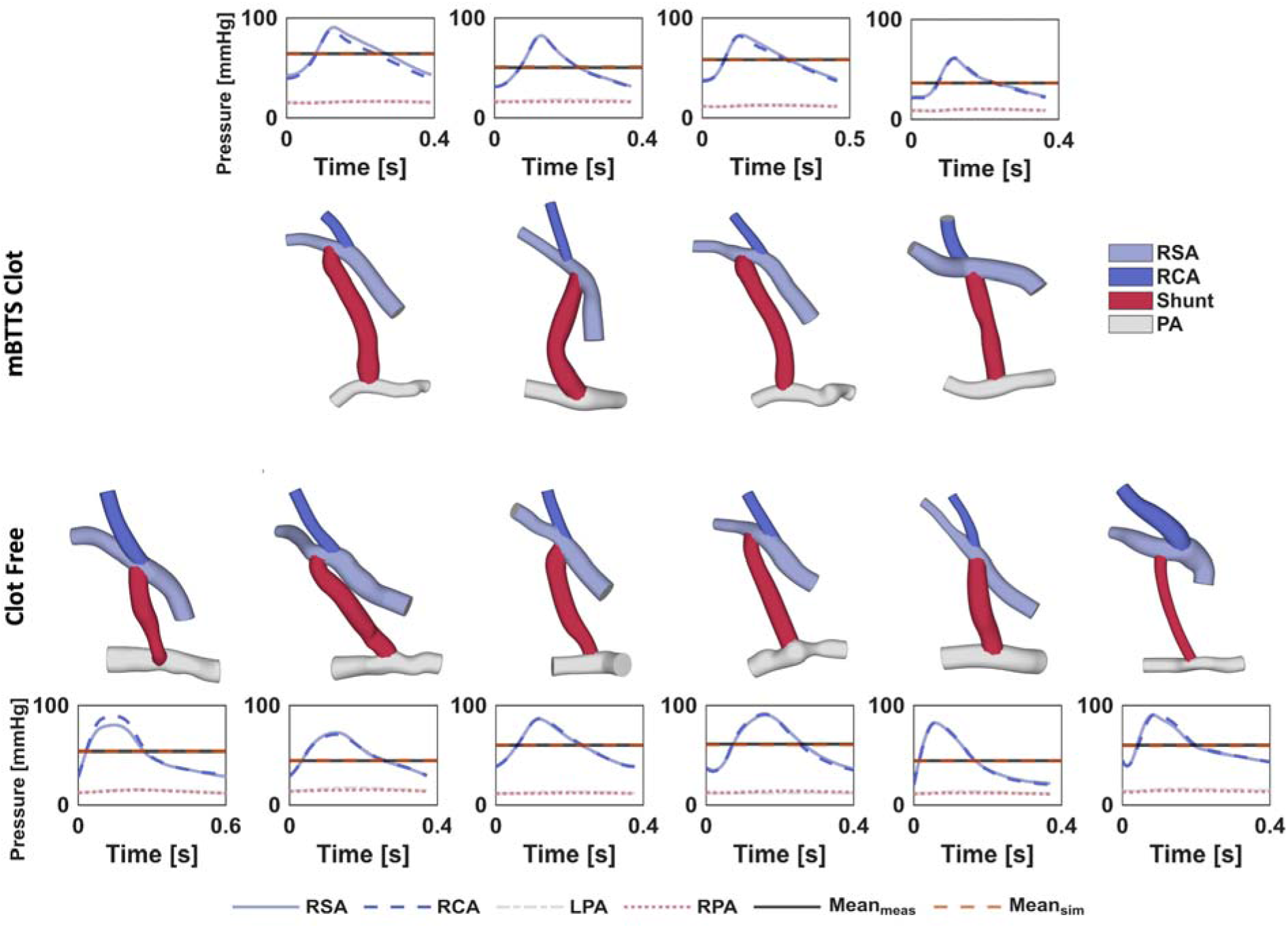
3D mBTTS Anatomic Geometries for All Patients. Three-dimensional reconstructions of mBTTS anatomies for all patients included in the study. Patients are grouped by clinical outcome: (top) mBTTS clot cases and (bottom) clot-free cases. The right subclavian artery (RSCA) is shown in light blue, the right carotid artery (RCA) in blue, the shunt graft in red, and the pulmonary artery (PA) in light gray. For each patient model, the corresponding simulated pressure waveforms at the RSCA, RCA, left PA (LPA), and right PA (RPA) outlets are shown. The solid gray horizontal line indicates the measured mean arterial pressure obtained from clinical catheterization data, while the dashed orange horizontal line indicates the cycle-averaged mean pressure from the simulation. Agreement between simulated and measured mean pressures confirms the physiologic consistency of the boundary conditions used in the simulations.

**Table 1.** Clinical Summary of mBTTS Patient Cohort (n=10)

| Pt . | Age at insert [days] | Shunt Size [mm] | Weight at insert [kg] | Sex | Type of CHD | Surgery | Unexpected Procedure | Shunt Thrombosis | Age at Clot [days] | Days to Clot [days] | Days to Shunt Removal or Death [days] | Antithrombotic | Associated Complications at Time of Clot | Clots Outside mBTTS | Survival |
| --- | --- | --- | --- | --- | --- | --- | --- | --- | --- | --- | --- | --- | --- | --- | --- |
| 1 | 1-10 | 3.5 | 2.76 | M | HLHS (MA/AA) | Norwood | — | Yes | 101-110 | 106 | 165 | ASA and Enoxaparin | Sepsis | IVC thrombus | Yes |
| 2 | 41-50 | 4 | 3.7 | F | HLHS (MS/AS) | Norwood | — | Yes | 91-100 | 53 | 216 | ASA | Sepsis | IVC thrombus | Yes |
| 3 | 1-10 | 3.5 | 4 | M | HLHS (MA/AA) | Norwood | mBTTS revision | Yes | 1-10 | 2 | 112 | Heparin | — | — | Yes |
| 4 | 1-10 | 3.5 | 3.265 | M | Situs inversus with dextrocardia, HLHS (MA/AA) | Norwood | — | Yes | 71-80 | 70 | 126 | ASA and Enoxaparin | — | — | Yes |
| 5 | 1-10 | 3.5 | 3.82 | M | HLHS (MA/AA) | Norwood | — | No | — | — | 119 | ASA | — | — | Yes |
| 6 | 1-10 | 3.5 | 3.97 | M | HLHS (MS/AA) | Norwood | — | No | — | — | 271 | ASA and Bivalirudin | — | Right Common femoral artery | No |
| 7 | 1-10 | 3.5 | 3.95 | F | HLHS with dextrocardia, PAPVR | Norwood | — | No | — | — | 270 | ASA | — | — | Yes |
| 8 | 1-10 | 3.5 | 4.1 | M | HLHS (MA/AA) | Norwood | — | No | — | — | 212 | ASA | — | — | Yes |
| 9 | 1-10 | 3.5 | 3.77 | F | HLHS (MA/AA) | Norwood | — | No | — | — | 190 | ASA | — | — | Yes |
| 10 | 1-10 | 3.5 | 3.15 | M | HLHS (MA/AA) | Norwood + TV repair | mBTTS balloon angioplasty + stenting | No | — | — | 145 | Bivalirudin | — | — | No |

\* HLHS= hypoplastic left heart syndrome; MA= mitral atresia; AA= aortic atresia; MS= mitral stenosis; AS= aortic stenosis; PAPVR= partial anomalous pulmonary venous return; TV= tricuspid valve; mBTTS= modified Blalock-Taussig-Thomas Shunt; ASA=aspirin; IVC= inferior vena cava.

### Boundary Conditions and Parameter Choice

In this study, physiologic pulsatile inflow was prescribed for all simulations. The inlet boundary condition was prescribed as a time-resolved volumetric flow rate waveform, derived from the velocity extracted via Doppler ultrasound. To distribute this flow, a flat velocity profile was applied, reflecting the underdeveloped flow typically observed due to the proximity to the aortic arch. For patients lacking Doppler velocity measurements at the innominate artery, the inlet waveform was reconstructed from Doppler data acquired at downstream vessels and adjusted using relative shunt velocity distributions based on the Conservation of Mass. The reconstructed inflow was then verified against cardiac catheterization measurements of flow rate and pressure to ensure physiologic consistency. To represent distal vascular impedance and capture non-periodic flow behavior, three-element Windkessel (RCR) models were applied at all outlets. The RCR model consists of a proximal resistance (R⍰) in series with a parallel combination of distal resistance (R_d_) and compliance (C).^29^ The RCR model was coupled to each outlet. The flow rate (Q) and area-averaged pressure (P) at each outlet are related by:

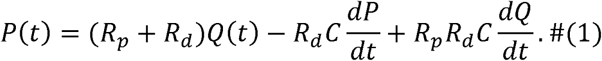

Resistance and capacitance values were individually tuned for each patient to match catheter-measured systolic, diastolic and mean pressures. The resulting simulated pressure waveforms at the RSCA, RCA, LPA, and RPA outlets were then compared with clinical data to confirm physiologic agreement. At each outflow boundary, a pressure Neumann boundary condition was imposed through the coupled RCR model. A backflow stabilization coefficient 0.5 was additionally applied to control flow reversal at outlets. Vessel walls were modeled as rigid with no-slip boundary conditions.

Geometric parameters found to be influential in our idealized study were systematically evaluated across patient-specific models to validate the previously observed trends: the distance between shunt insertion and right carotid artery (RCA) insertion (L_1_), the angle between the shunt and pulmonary artery (*α*), the pulmonary artery diameter (D_PA_) and shunt diameter (D_shunt_).

### Mesh Generation and Validation

Surface and volume meshes were generated using SimVascular (**Figure 1B**). To resolve near-wall flow, eight prismatic boundary layers were applied along vessel walls, while the remaining volume was discretized with tetrahedral elements. Inflation layers were generated using a relative thickness of 0.25 edge length and a growth rate of 1.1, ensuring adequate resolution of boundary layer gradients, important to accurately quantifying shear stress. A mesh convergence study was performed to balance accuracy and computational cost. The solution converged at approximately 560,000 elements, beyond which further mesh refinement produced less than 1.8% change in peak of time-averaged wall shear stress (Peak WSS). Therefore, all patient-specific models were meshed with approximately 600,000 elements, with minor adjustments to account for anatomical variation (**Supplemental Figure 1**). Because solution periodicity is influenced by the RCR outlet models, simulations were continued until a periodic state was reached. All quantitative results were extracted from the final cardiac cycle.

### Numerical Simulations and Post-Processing

Transient simulations were performed by solving the three-dimensional, time-dependent Navier-Stokes equations using the finite-element solver svMultiPhysics (SimVascular)^26,30^. A time step of 0.0008-0.001 s was used, with 3-40 nonlinear iterations per step to ensure convergence of the residual at every time step. Blood was modeled as an incompressible fluid with a density of 1060 kg/m^3^, and its shear-thinning behavior was represented using the Carreau-Yasuda viscosity model, with parameters μ_0_ = 22 × 10^-3^ Pa · s, μ_∞_ = 2.2 × 10^-3^ Pa · s, a = 0.644, n = 0.392, λ = 0.110 s.^31^ Inlet Reynolds number across the cohort ranged from Re=1566-2225, and peak systolic Re remained below 2800 in all cases. Although these values approach the classical pipe-flow transition threshold, prior computational and experimental studies of systemic-to-pulmonary shunts and comparable small-vessel anastomoses have demonstrated that laminar modeling adequately captures the dominant flow features in this regime, with transitional effects confined to small, localized regions near anastomotic junctions.^32–35^ A laminar governing model was applied for all simulations. Note that while this is adequate for predicting the onset of thrombosis, turbulence can be expected in diseased vessels undergoing stenosis;^3^ the potential influence of localized transitional flow is addressed in the Discussion.

For each patient model, two hemodynamic parameters associated with thrombosis risk, WSR and ESR, were computed. Time-resolved simulation results from the last cardiac cycle were extracted and processed in ParaView (Clifton Park, NY). Temporal statistics were calculated over the full cycle to obtain peak and mean values, and systolic and diastolic time steps were extracted at their corresponding time step to evaluate instantaneous WSR and ESR at each time point. These processed data were then used to quantify spatial distributions and to examine the relationship between local hemodynamics with shunt geometry.

WSR was calculated as:

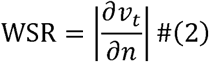

where *v_t_* is velocity component parallel to the wall, and *n* is the direction perpendicular to the wall. ESR, quantifying the rate of stretching along flow streamlines, was computed as the normal component of the strain rate tensor projected along the local velocity direction:

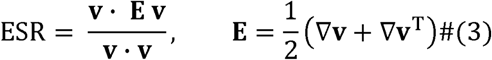

Where **v** is the local velocity vector and **E** is the strain rate tensor.

To enable comparison across patients with varying cardiac outputs, both metrics were normalized by characteristic values derived from the mean inlet flow rate (*Q_m_*) and inlet diameter (*D_in_*):

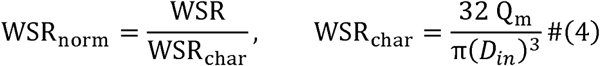

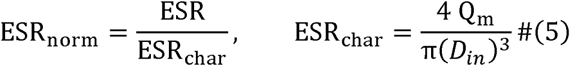

The characteristic values represent the theoretical wall shear and convective acceleration at the inlet, respectively. Normalizing by these values ensures that the resulting dimensionless parameters isolate the influence of shunt geometry from absolute variations in physiologic flow magnitude.

To minimize the influence of localized numerical artifacts, particularly near sharp geometric corners or regions of mesh singularity, peak hemodynamic values were not extracted from single surface or volume elements. Instead, they were computed over localized regions identified via thresholding to minimize sensitivity to pointwise numerical discrepancies that are unlikely to exist *in vivo*. Because regions of elevated hemodynamic stress are spatially distributed, identifying a single maximum value can be sensitive to these localized artifacts. Therefore, in addition to reporting peak values, threshold metrics were introduced.

For WSR, two thresholds were applied to characterize the stages of thrombus formation. A primary threshold of 5,000 s^−1^ was used to identify regions where von Willebrand Factor (VWF) typically begins to unfold from its globular state into an elongated conformation, a prerequisite for platelet adhesion^21^. A secondary threshold of 20,000 s^−1^ was applied to identify higher-risk regions where shear-induced platelet aggregation becomes more prevalent^22^. At this level, VWF-platelet aggregates have been shown to occur independently of activation. Similarly, for ESR, a threshold of 400 s^−1^ was applied to calculate the fraction of the fluid domain exceeding this value relative to the total volume^23^. This threshold has been theorized to trigger VWF extension and the unfolding of the VWF A1 domain that can trigger platelet glycoprotein (GP) Ib-IX signaling that leads to granule release and can ultimately trigger shear-induced platelet activation.^36^

### Statistics

Associations between geometric parameters and hemodynamic metrics were evaluated using Spearman’s rank correlation coefficient (*ρ*), which quantifies the strength and direction of monotonic relationships between variables. Linear regression lines were included in scatter plots to illustrate trends between variables. Correlations were assessed between shunt insertion angle deviation, shunt diameter, and key hemodynamic metrics. Given the cohort size, these analyses were designed to identify geometric trends, with statistical significance interpreted as supportive rather than definitive. Statistical analyses and figure generation were performed in MATLAB (MathWorks, Natick, MA). A p-value < 0.05 was considered statistically significant.

### Data Sharing Statement

The data supporting the findings of this study are available from the corresponding author upon reasonable request and subject to institutional and ethical restrictions.

## RESULTS

### Simulated Pressures Match Clinical Measurements

Simulated systolic, diastolic, and mean pressures at all four outlets (RSCA, RCA, LPA, RPA) showed close agreement with catheter-derived clinical measurements across all 10 patient-specific models (**Figure 2**). Calibrated resistance and capacitance values for each outlet are listed in **Table 2**. This correspondence confirms that the patient-specific RCR boundary conditions adequately capture downstream vascular impedance and provide a physiologically grounded foundation for the hemodynamic analyses that follow.

**Table 2:** Resistance and Capacitance Values for RCR boundary conditions for each model.

| Pt. | RSCA |  |  | RCA |  |  | LPA |  |  | RPA |  |  |
| --- | --- | --- | --- | --- | --- | --- | --- | --- | --- | --- | --- | --- |
|  | Rp | Rd | C | Rp | Rd | C | Rp | Rd | C | Rp | Rd | C |
| 1 | 5210 | 7430 | 0.81 | 9310 | 21900 | 0.94 | 228 | 4010 | 9.5 | 143 | 3040 | 11.0 |
| 2 | 3820 | 5740 | 0.39 | 8540 | 12900 | 0.53 | 167 | 3340 | 22.0 | 443 | 6320 | 17.0 |
| 3 | 3250 | 6970 | 0.76 | 7510 | 17500 | 0.9 | 298 | 3730 | 8.3 | 187 | 2340 | 9.5 |
| 4 | 319 | 4240 | 0.23 | 3400 | 5000 | 0.77 | 189 | 2370 | 9.3 | 139 | 1990 | 10.4 |
| 5 | 4380 | 9920 | 1.8 | 2040 | 9190 | 7.9 | 91 | 1290 | 18.0 | 137 | 1710 | 17.0 |
| 6 | 5870 | 7940 | 1.5 | 6300 | 8990 | 1.33 | 272 | 2200 | 9.8 | 633 | 4520 | 6.5 |
| 7 | 5240 | 8580 | 0.38 | 8020 | 13600 | 0.45 | 81 | 1400 | 16.8 | 344 | 3820 | 9.0 |
| 8 | 3700 | 9030 | 1.6 | 4150 | 11200 | 1.87 | 245 | 3770 | 20.0 | 166 | 2210 | 19.5 |
| 9 | 5660 | 7540 | 0.19 | 7740 | 11100 | 0.3 | 93 | 1860 | 21.0 | 356 | 3960 | 16.0 |
| 10 | 8490 | 17500 | 2.3 | 3210 | 10200 | 3.95 | 110 | 2620 | 15.3 | 690 | 5110 | 6.3 |

\* Rp is the proximal resistance, dyn s/cm<sup>5</sup>, Rd is the distal resistance, dyn s/cm<sup>5</sup>, C is the capacitance, × 10<sup>-5</sup>cm<sup>5</sup>/ dyn.

### Peak Shear Metrics Localize to the Shunt–Subclavian Junction

Despite substantial interpatient anatomic variability, elevated WSR and ESR consistently localized to the shunt insertion region across all 10 patient-specific models (**Figure 3**). Peak time-averaged WSR values occurred at the shunt wall immediately distal to the shunt–subclavian artery junction in 8 of 10 patients, including all 4 thrombosed cases and 4 of the 6 patent cases; the remaining 2 patent cases showed peaks at the shunt-PA anastomosis. Peak ESR demonstrated a nearly identical spatial pattern (**Figure 3C**) with 8 of 10 patients exhibiting maxima at the shunt-subclavian bifurcation. No peak values for either metric were observed within the distal RSCA or RCA branches.

**Figure 3:**
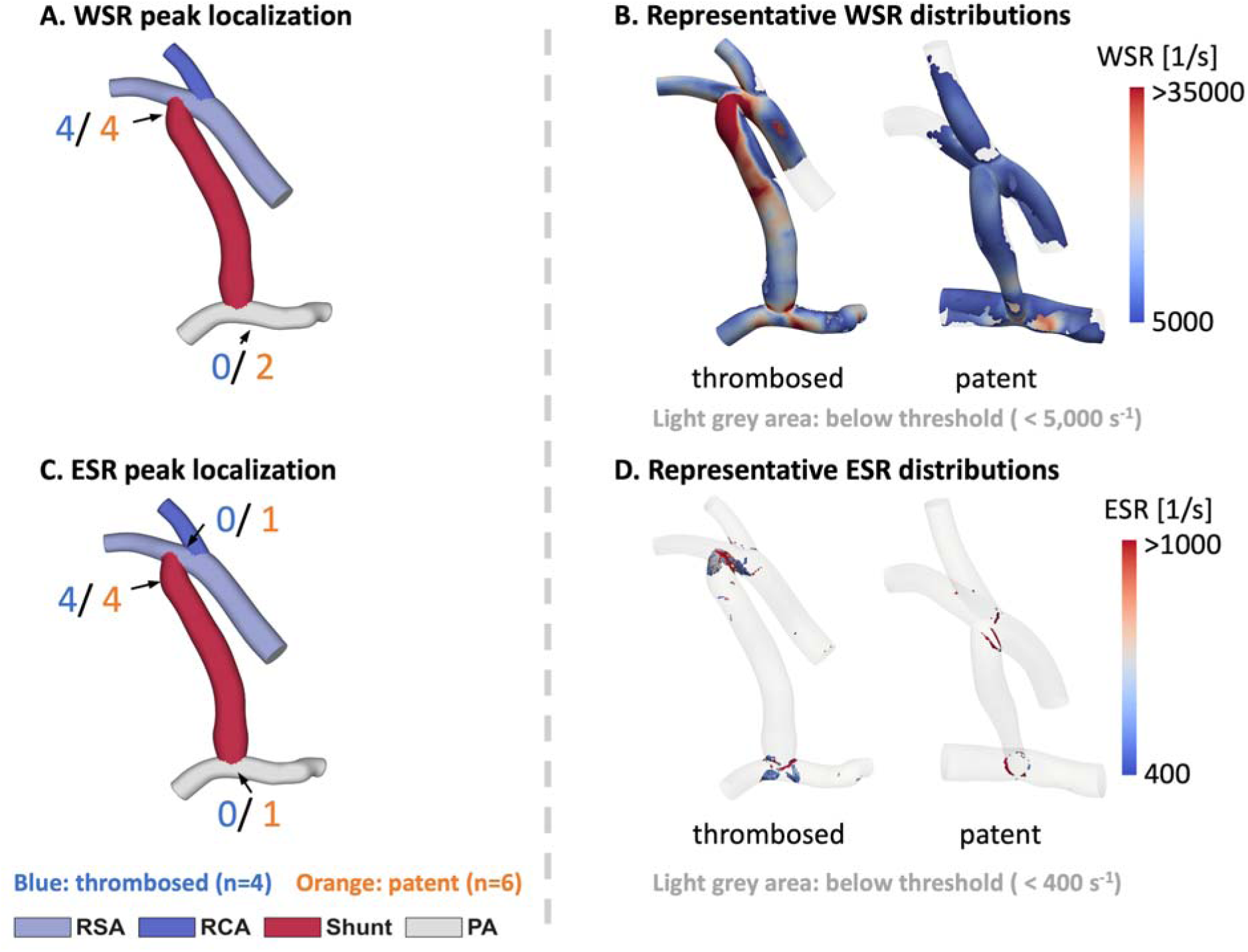
Spatial Distribution of Peak Hemodynamic Metrics and Representative Examples. **Left panels** (A, C) **show** frequency maps of peak time-averaged wall shear rate (WSR) and elongational strain rate (ESR) localization across all 10 patient-specific models; frequency counts indicate the number of patients from each cohort (thrombosed: n=4, blue; patent: n=6, orange) in whom the peak value occurred at that anatomic location (e.g., “4/4”). Right panels (**B, D**) show representative time-averaged WSR (threshold: > 5,000 s^-1^) and ESR (threshold: > 400 s^-1^) surface distributions for one thrombosed and one patent case; light grey regions fall below the respective threshold. In 8 of 10 patients, peak WSR and ESR localized to the shunt immediately distal to the shunt-subclavian artery junction.

The spatial distribution of high wall shear exposure across individual vessels is summarized in **Table 3**. The VWF-unfolding threshold (WSR > 5,000 s⁻¹) was exceeded broadly across all four vessels. At the higher platelet-aggregation threshold (WSR > 20,000 s⁻¹), however, the RCA consistently contributed the smallest share of high-shear surface area across the cohort (<1.4% of total model surface area), whereas the RSCA and shunt together accounted for the majority of the high-shear area (**Table 3**), further supporting localization of extreme shear to the shunt–RSCA junction. The only exceptions were the two patent cases in which the shunt–PA anastomosis contributed the largest fraction of high-shear surface area, consistent with the peak WSR localization in those cases. Representative WSR and ESR distributions for a thrombosed and patent case (**Figure 3B, D**) highlight that the spatial distribution of elevated stress appears more extensive in the thrombosed cohort. This localization pattern is consistent with the 70% rate of peak shear concentration at the shunt-subclavian bifurcation observed in a prior study of idealized configurations.^24^

**Table 3:**
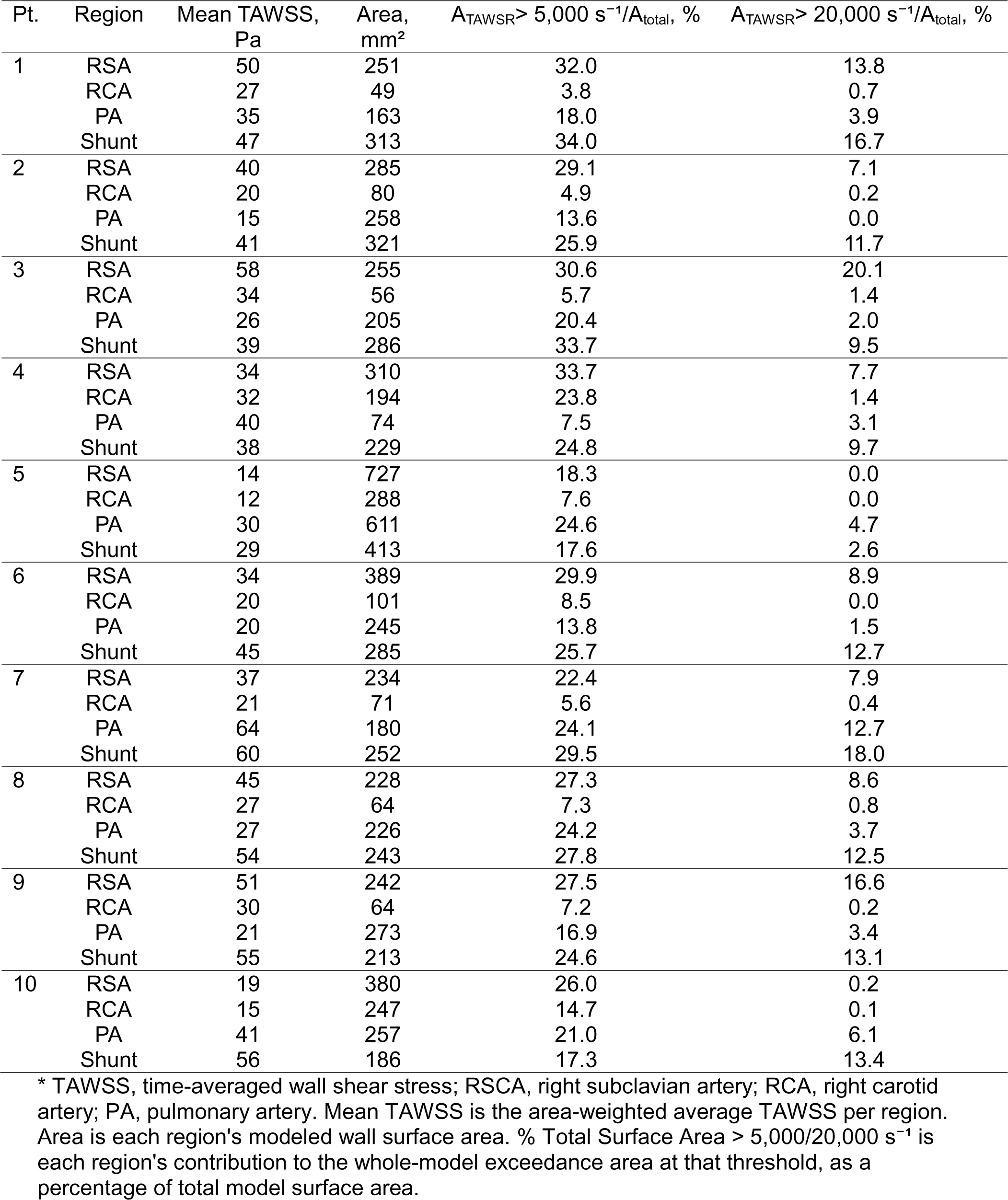
Regional Distribution of High-Shear Exposure Across the Patient-Specific Models.

| Pt. | Region | Mean TAWSS, Pa | Area, mm<sup>2</sup> | $A_{TAWSR > 5,000 \text{ s}^{-1}/A_{\text{total}}, \%}$ | $A_{TAWSR > 20,000 \text{ s}^{-1}/A_{\text{total}}, \%}$ |
| --- | --- | --- | --- | --- | --- |
| 1 | RSA | 50 | 251 | 32.0 | 13.8 |
|  | RCA | 27 | 49 | 3.8 | 0.7 |
|  | PA | 35 | 163 | 18.0 | 3.9 |
|  | Shunt | 47 | 313 | 34.0 | 16.7 |
| 2 | RSA | 40 | 285 | 29.1 | 7.1 |
|  | RCA | 20 | 80 | 4.9 | 0.2 |
|  | PA | 15 | 258 | 13.6 | 0.0 |
|  | Shunt | 41 | 321 | 25.9 | 11.7 |
| 3 | RSA | 58 | 255 | 30.6 | 20.1 |
|  | RCA | 34 | 56 | 5.7 | 1.4 |
|  | PA | 26 | 205 | 20.4 | 2.0 |
|  | Shunt | 39 | 286 | 33.7 | 9.5 |
| 4 | RSA | 34 | 310 | 33.7 | 7.7 |
|  | RCA | 32 | 194 | 23.8 | 1.4 |
|  | PA | 40 | 74 | 7.5 | 3.1 |
|  | Shunt | 38 | 229 | 24.8 | 9.7 |
| 5 | RSA | 14 | 727 | 18.3 | 0.0 |
|  | RCA | 12 | 288 | 7.6 | 0.0 |
|  | PA | 30 | 611 | 24.6 | 4.7 |
|  | Shunt | 29 | 413 | 17.6 | 2.6 |
| 6 | RSA | 34 | 389 | 29.9 | 8.9 |
|  | RCA | 20 | 101 | 8.5 | 0.0 |
|  | PA | 20 | 245 | 13.8 | 1.5 |
|  | Shunt | 45 | 285 | 25.7 | 12.7 |
| 7 | RSA | 37 | 234 | 22.4 | 7.9 |
|  | RCA | 21 | 71 | 5.6 | 0.4 |
|  | PA | 64 | 180 | 24.1 | 12.7 |
|  | Shunt | 60 | 252 | 29.5 | 18.0 |
| 8 | RSA | 45 | 228 | 27.3 | 8.6 |
|  | RCA | 27 | 64 | 7.3 | 0.8 |
|  | PA | 27 | 226 | 24.2 | 3.7 |
|  | Shunt | 54 | 243 | 27.8 | 12.5 |
| 9 | RSA | 51 | 242 | 27.5 | 16.6 |
|  | RCA | 30 | 64 | 7.2 | 0.2 |
|  | PA | 21 | 273 | 16.9 | 3.4 |
|  | Shunt | 55 | 213 | 24.6 | 13.1 |
| 10 | RSA | 19 | 380 | 26.0 | 0.2 |
|  | RCA | 15 | 247 | 14.7 | 0.1 |
|  | PA | 41 | 257 | 21.0 | 6.1 |
|  | Shunt | 56 | 186 | 17.3 | 13.4 |

\* TAWSS, time-averaged wall shear stress; RSA, right subclavian artery; RCA, right carotid artery; PA, pulmonary artery. Mean TAWSS is the area-weighted average TAWSS per region. Area is each region's modeled wall surface area. % Total Surface Area $> 5,000/20,000 \text{ s}^{-1}$ is each region's contribution to the whole-model exceedance area at that threshold, as a percentage of total model surface area.

### Shunt Insertion Angle and Diameter Are Primary Geometric Determinants of Hemodynamic Stress

Shunt insertion angles across the cohort ranged from 66° to 138° (median 94°), and shunt internal diameters at the time of CT imaging ranged from 2.3 to 3.9 mm (median 3.6 mm) (**Figure 4**). To isolate the influence of graft geometry from variations in patient cardiac output, all hemodynamic metrics were normalized by patient-specific inlet conditions.

**Figure 4:**
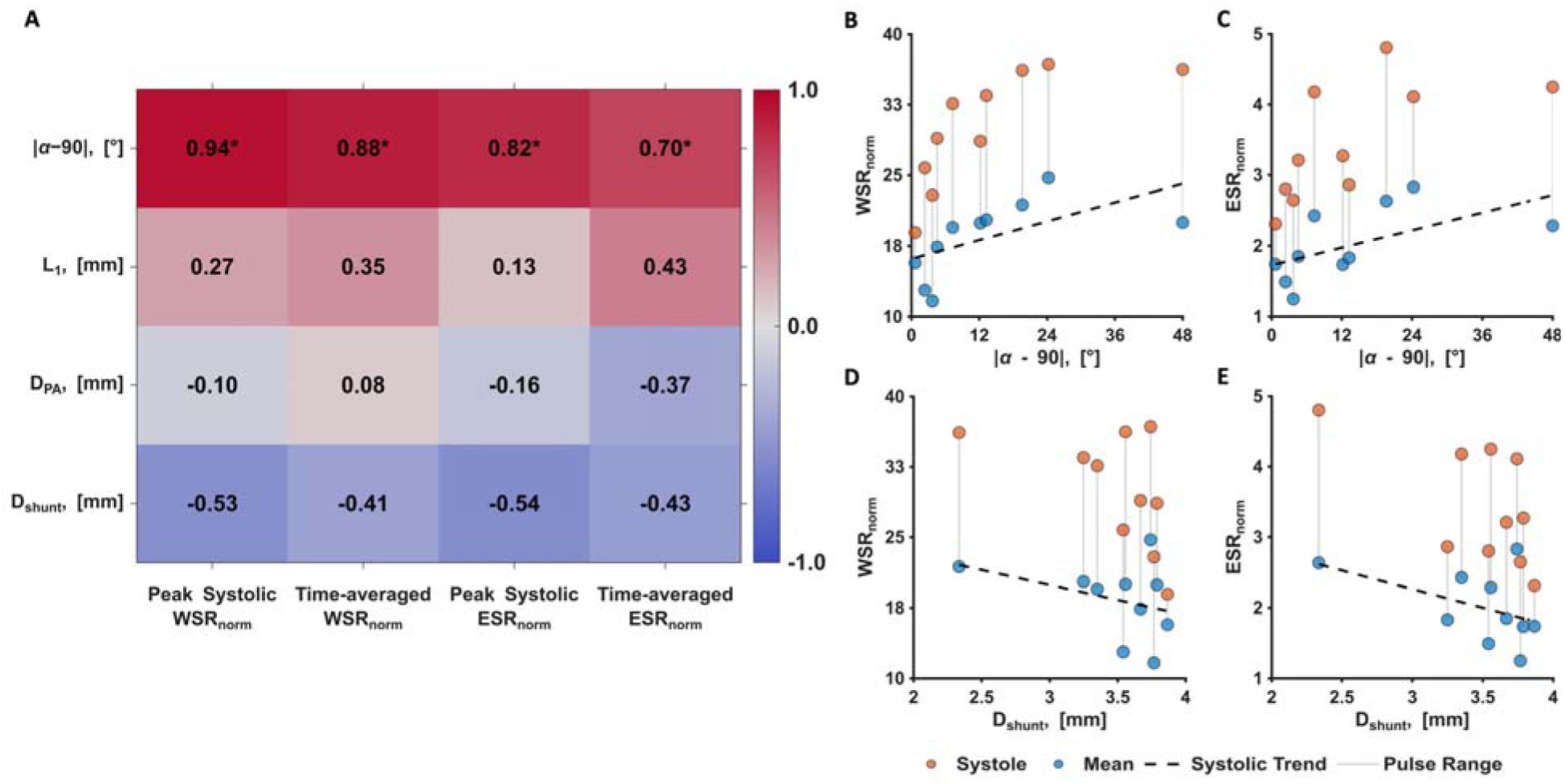
Geometric Influence on Hemodynamic Metrics in Patient-specific mBTTS Models. **A,** Heatmap of Spearman correlation coefficients (*ρ*) illustrating the relationship between shunt geometric parameters (L_1_, α, D_shunt_, and D_PA_) and normalized hemodynamic metrics (WSR_norm_ and ESR_norm_) at peak systole and time-averaged states. Statistical significance (p<0.05) is denoted by asterisks (*). The parameter (|α − 90°|) represents the absolute deviation from a perpendicular (90°) shunt insertion, e.g. both 80° and 100° corresponding to a 10° deviation. **B-E,** Scatter plots of primary geometric drivers. A unified legend at the bottom defines the visual encoding: orange markers represent peak systolic values, blue markers represent time-averaged values, and vertical gray lines connect these values for each patient model. Dashed lines indicate linear regression fits for the *time-averaged* data. **Angle Deviation:** Significant correlations were observed between |α − 90°| and the time-averaged metrics (p < 0.05). **Shunt diameter**: An inverse relationship was noted for D_shunt_ across all metrics, suggesting that larger diameters generally attenuate peak stressors, though these trends approached but did not reach the p < 0.05 threshold in this cohort.

Correlations between four geometric parameters (L_1_, α, D_shunt_, and D_PA_) and normalized WSR and ESR at both peak systole and cycle-averaged time points (**Figure 4A**) show that insertion angle emerged as the strongest geometric predictor of hemodynamic stress. The absolute deviation of the insertion angle from perpendicular (|α −90°|) correlated significantly with both systolic and time-averaged WSR_norm_ (*ρ* = 0.94, *p* = 0; *ρ* = 0.88, *p* = 0.002) and ESR_norm_ (*ρ* = 0.82, *p* = 0.007; *ρ* = 0.70, *p* = 0.03) (**Figure 4B,C**). Configurations closer to perpendicular insertion were associated with lower shear exposure, whereas increasingly oblique insertion angles corresponded to progressively elevated wall shear and elongational strain at the anastomosis.

Shunt diameter showed a concordant inverse relationship with hemodynamic stress across all four metrics. Larger graft diameters were associated with lower time-averaged WSR_norm_ (*ρ* = −0.41, *p* = 0.25) and ESR_norm_ (*ρ* = −0.43, *p* = 0.22), with similar trends at peak systole (*ρ* = −0.53 and *ρ* = −0.54, respectively) (**Figure 4D, E**). Although these associations did not reach significance in this cohort, the consistent directionality across all metrics and time points suggests an underlying trend.

Neither the distance between shunt and right carotid artery insertions (*L*_1_) nor pulmonary artery diameter (*D_PA_*) showed meaningful associations with any hemodynamic metric (**Figure 4A**), indicating that among the geometric parameters evaluated, insertion angle and shunt diameter are the primary surgically modifiable determinants of local shear exposure.

### Thrombosed Shunts Exhibit Elevated Cycle-Averaged Shear Exposure

Hemodynamic metrics and threshold-based exposure fractions were compared between thrombosed (*n* = 4) and patent (*n* = 6) shunts (**Figure 5**). Given the cohort size, these comparisons were descriptive and intended to assess directional consistency with the geometric findings rather than to establish formal statistical differences.

**Figure 5:**
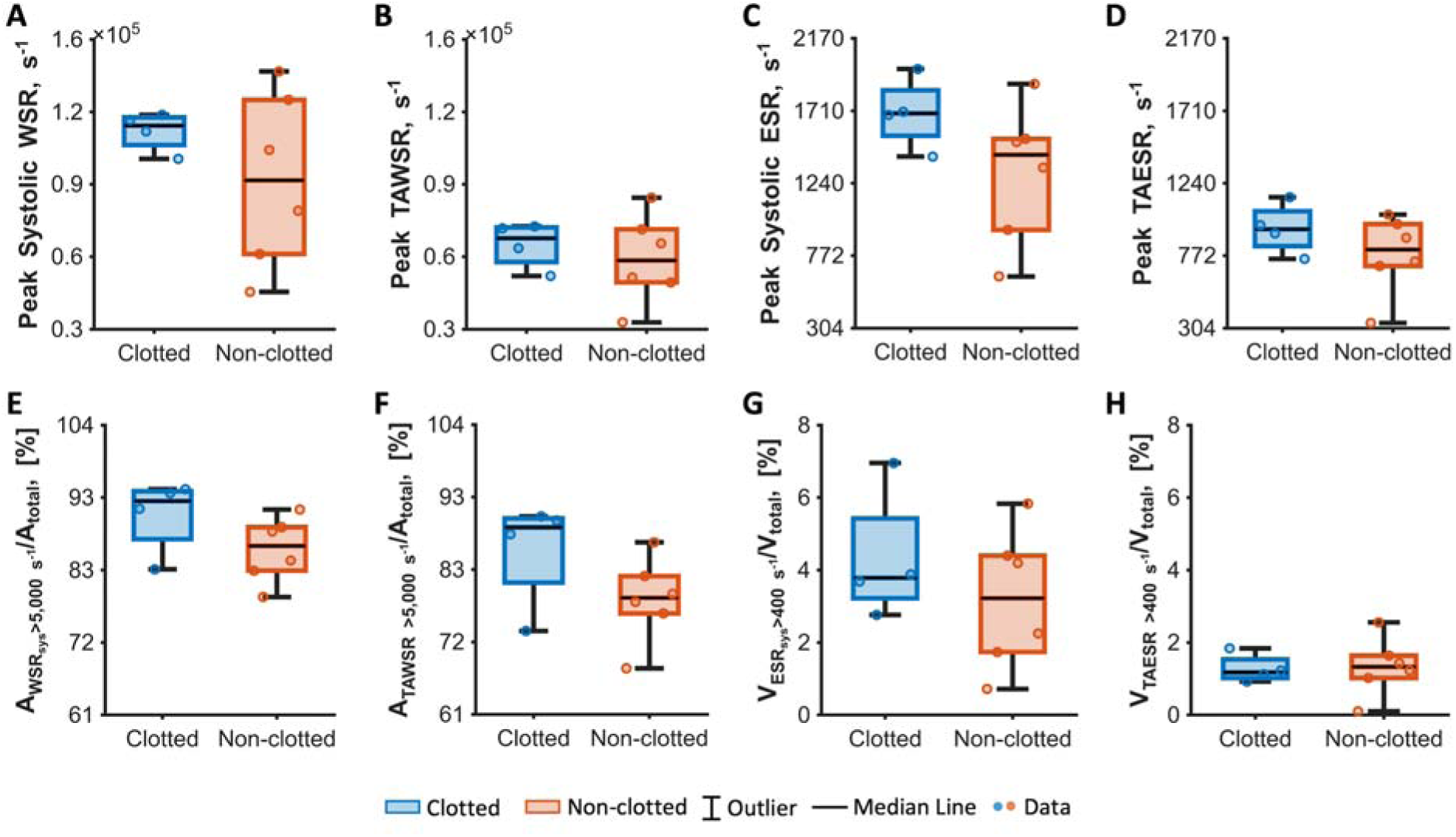
Comparison of Hemodynamic Metrics between mBTTS Clot and Clot-free Groups. *Box plots* illustrate peak systolic and time-averaged flow metrics (wall shear rate [WSR] and elongational strain rate [ESR]) for clotted (n=4, blue) and clot-free (n=6, orange) groups. *Boxes* display median, quartiles, and range for each group. **A,** Peak WSR at systole. **B,** Peak TAWSR. **C,** Peak ESR at systole. **D,** Peak TAESR. **E,** Surface area fraction with WSR > 5,000 s^−1^ (A_WSR_ > 5,000 s^−1^/ A_total_) at systole. **F,** Surface area fraction with time-averaged WSR > 5,000 s^−1^ (A_WSR_ > 5,000 s^−1^/ A_total_). **G,** Volume fraction with ESR>400 s^−1^ (V_ESR_>400 s^−1^/V_total_) at systole. **H,** Time-averaged volume fraction with ESR>400 s^−1^ (V_TAESR_>400 s^−1^/V_total_). A_total_ and V_total_ represent the total surface area and total volume of the fluid domain, respectively. (25th-75th quartiles: A, 1.1 – 1.2 × 10^5^, 0.6 – 1.3 × 10^5^. B, 5.8 – 7.3 × 10^4^, 4.9 – 7.2 × 10^4^. C, 1,540-1,840, 938-1,520. D, 830-1,060, 705-976. E, 87.2%-94.1%, 82.5%-89.0%. F, 80.0%-90.0%, 76.1%-81.6%. G, 3.2%-5.3%, 1.7%-4.3%. H, 1.0%-1.5%, 1.0%-1.6%).

At peak systole, WSR distributions overlapped between groups, with wider spread in the patent group (IQR 1.1 – 1.2 × 10^5^ vs. 0.6 – 1.3 × 10^5^, **Figure 5A**). When averaged over the cardiac cycle, the two groups showed less overlap between their interquartile ranges, with a higher median peak TAWSR in the thrombosed group (median 6.8 × 10^4^, IQR 5.8 – 7.3 × 10^4^ vs. 4.9 – 7.2 × 10^4^, **Figure 5B**). ESR followed the same trend, with thrombosed cases exhibiting higher values at both peak systole (median 1,690, IQR 1,540-1,840 vs. 938-1,520, **Figure 5C**) and cycle-averaged time points (median 943, IQR 830-1,060 vs. 705-976, **Figure 5D**).

Threshold-based exposure metrics showed a similar pattern. The surface area fraction exceeding the VWF-unfolding threshold of 5,000 s^−1^ was higher in the thrombosed group at both peak systole (median 92.7%, IQR 87.2-94.1% vs. 82.5-89.0%, **Figure 5E**) and when cycle-averaged (median 88.8%, IQR 80.0-90.0% vs. 76.1-81.6%, **Figure 5F**). The volumetric fraction exceeding the ESR threshold of 400 s⁻¹ was greater in the thrombosed group at peak systole (median 3.7% vs. 3.1%, IQR 3.2-5.3% vs. 1.7-4.3%, **Figure 5G**), although this separation did not persist when cycle-averaged. Surface area fractions exceeding the platelet aggregation threshold of 20,000 s⁻¹ showed a similar directional trend that was most evident in the cycle-averaged comparison (median 27.4% vs. 24.3%) (**Supplemental Figure 2**).

Across both peak and threshold-based metrics, peak systolic values generally demonstrated equal or greater separation between thrombosed and patent shunts compared with cycle-averaged values, suggesting that transient systolic shear extremes may provide sensitive indicators of thrombogenic flow environments.

## DISCUSSION

In this patient-specific computational study, we demonstrate that surgically modifiable geometric features of the mBTTS shunt remain key determinants of thrombogenic hemodynamics in anatomically realistic configuration. After calibrating boundary conditions against catheter-derived pressure measurements, we identified three central findings. First, peak WSR and ESR consistently localized to the shunt-subclavian junction across anatomically-diverse patients, establishing this region as a key biomechanical vulnerability. Second, deviation of the shunt Insertion angle from perpendicular correlated strongly with local shear exposure, identifying insertion angle as a primary surgically modifiable determinant of thrombogenic flow conditions. Finally, shunt diameter showed a concordant inverse relationship with shear metrics, and thrombosed shunts exhibited higher shear exposure across several peak and cycle-averaged measures than patent shunts, suggesting a possible mechanistic link between geometric design and clinical thrombosis. Collectively, these findings provide translational validation that principles identified in idealized computational models persist *in vivo* and motivate the integration of hemodynamic assessment into precision surgical planning.

### A Reproducible Biomechanical Vulnerability at the Shunt–Subclavian Junction

Peak shear metrics consistently localized to a single anatomic region, the shunt wall immediately distal to the shunt–subclavian junction, despite substantial variation in overall anatomy, insertion position, and pulmonary artery configuration. This spatial reproducibility, observed in 8 of 10 patients and in all 4 thrombosed cases, closely parallels our prior idealized modeling results and indicates that thrombosis risk at the mBTTS is not distributed randomly across the conduit but arises from a predictable flow disturbance at the proximal anastomosis.^24^ The mechanistic basis is well established: abrupt geometric transitions generate localized jet impingement and elevated wall shear, conditions that unfold von Willebrand factor, expose its platelet-binding A1 domain, and initiate shear-mediated platelet adhesion and aggregation.^9,10,14,17,37,38^ What this study adds is the observation that this biomechanical vulnerability is reproducible across patient-specific anatomies and preserved under physiologic pulsatile flow, strengthening the biological plausibility of geometry as a modifiable risk factor and focusing attention on a specific region of the shunt for preoperative optimization.

The magnitude of shear exposure predicted by our models is high, but is corroborated by the clinical hematologic phenotype already documented in this patient population. First, acquired von Willebrand syndrome (aVWS) is ubiquitous with mBTTS patients, which is a process understood to be caused by shear-induced VWF unfolding and ADAMTS13-mediated proteolysis of high-molecular-weight VWF multimers, requiring high shear rates typically above 5000 s^-1^.^39–41^ Hemolysis, which requires substantially higher and more sustained shear exposure, is less uniformly observed but remains a recognized complication, documented in case reports of hemolytic anemia following mBTTS placement and in independent computational modeling of comparable Norwood geometries, which report peak wall shear rates in the same 10^4^-10^5^ s^-1^ range identified in the present study alongside a clinically significant post-operative hematocrit decline.^42,43^ Therefore, the clinical outcomes for these patients support the biological plausibility of the shear magnitudes we report in the current study, which are very high relative to reported physiological values.

### Insertion Angle and Diameter as Surgically Actionable Targets

The significant association between more oblique insertion angles and increased time-averaged normalized wall shear rate, combined with the inverse trend for shunt diameter, identifies two geometric parameters that surgeons can modify at the time of implantation. Oblique insertions produce asymmetric jet impingement and intensified near-wall velocity gradients at the anastomosis, while smaller shunt diameters increase bulk velocity and shear magnitude for a given flow rate. These trends held across anatomically diverse patient-specific models, suggesting that these geometric effects are not artifacts of idealization but reflect persistent flow physics *in vivo*.

These findings do not imply that a single optimal geometry exists for all patients. Insertion angle and diameter must be balanced against anatomic feasibility, systemic-to-pulmonary flow ratio, and pulmonary vascular resistance, all of which constrain surgical decision-making in ways that uniform shear minimization cannot capture. Rather, the results support a framework in which hemodynamic consequences of candidate configurations can be quantitatively evaluated alongside traditional surgical considerations, enabling individualized optimization within the constraints of a given patient’s anatomy.

#### Peak Systolic Hemodynamics Capture Key Thrombogenic Flow Features

A secondary observation of potential clinical significance is that thrombosed shunts were more clearly distinguished from patent shunts by peak systolic shear metrics than by cycle-averaged measures. Although the small cohort precludes formal statistical inference, this pattern was consistent across both shear magnitudes and threshold-based exposure fractions, consistent with prior CFD studies of comparable aorta-pulmonary shunt configurations, which have similarly emphasized peak systolic wall shear as the primary thrombogenic exposure metric.^32,44^

Platelet activation is governed by the interplay between shear magnitude and exposure duration, with sufficiently high shear capable of inducing rapid platelet activation even during brief exposure.^9,10^ Likewise, VWF undergoes force-dependent conformational changes under elevated shear, promoting platelet adhesion and thrombus initiation in regions of localized flow acceleration.^45,46^ In highly pulsatile shunt flows, transient systolic shear extremes may therefore be particularly relevant to these early thrombogenic processes. If confirmed in larger cohorts, this observation would support incorporating peak systolic and exposure-based measures in CFD-based surgical planning and thrombosis risk assessment.

### Clinical Translation: Shifting from Pharmacological to Geometrical Intervention

mBTTS thrombosis occurs despite systemic anticoagulation that places these infants among the most heavily anticoagulated patients in pediatric cardiology. The persistence of thrombotic events under aggressive pharmacologic management reflects a fundamental mismatch between the mechanism of therapy and the mechanism of disease: anticoagulants target circulating coagulation factors and platelet reactivity, but the trigger for thrombosis at the shunt is a local mechanical environment that anticoagulation does not modify. A central implication of this work is that mBTTS thrombosis risk may be partially modifiable by surgeon control of geometry at the time of surgical implantation.

In contrast to fixed patient-specific anatomy, insertion angle and graft diameter represent intraoperative decisions. Our prior idealized modeling suggested that distal positioning and near-perpendicular insertion with adequate diameter minimized adverse hemodynamic stressors, which would have to be balanced against controlling the systemic-to-pulmonary flow ratio. The present patient-specific validation demonstrates that these geometric principles persist despite anatomic variability and pulsatile physiologic flow.

This raises the possibility of incorporating preoperative imaging and computational modeling into surgical planning workflows. Patient-specific simulation could enable evaluation of candidate graft configurations before implantation, thereby selecting geometries that reduce shear-related exposure while preserving adequate pulmonary blood flow. Such an approach aligns with broader efforts toward precision congenital heart surgery, in which biomechanics informs operative strategy.

Importantly, these findings do not argue against anticoagulation therapy but instead suggest that pharmacologic and geometric strategies may be synergistic. By reducing local mechanical triggers, optimized graft design may lower the threshold for platelet activation and propagation of the coagulation cascade, potentially decreasing dependence on intensified systemic anticoagulation in vulnerable infants. Notably, any shear-induced platelet activation could lead to the release of activation agonist and additional platelet signaling that can help be mitigated by antiplatelet therapies. Anticoagulants could help prevent the coagulation cascade from progressing, which would otherwise stabilize primary thrombosis resulting from mechanical stimuli. Therefore, a more robust approach that considers hemodynamics could improve patient outcomes, supplementing a thrombotic pathway not currently targeted.

### Limitations and Future Directions

This study has several limitations. First, the cohort size was modest and retrospective, limiting statistical power and generalizability. Although patient-specific anatomies and physiologic boundary conditions were used, the observed associations should be considered hypothesis-generating and require validation in larger prospective cohorts.

Second, modeling assumptions may influence absolute shear estimates. Vessel walls were treated as rigid, and a laminar flow model was applied based on inlet Reynolds number analysis. While appropriate for global flow conditions, localized transitional or disturbed flow near anastomoses may not be fully captured. Incorporation of compliant wall modeling and advanced flow modeling approaches may further refine characterization of thrombogenic stress. Furthermore, shear values are extremely high compared to reported values, which are mostly for adult studies. Given the limits in imaging resolution and segmentation, a small misestimation of size could have a large impact on shear.

Third, thrombosis is multifactorial. Patient-specific hematologic factors, anticoagulation regimens, endothelial response, and postoperative hemodynamic adaptation were not incorporated into the simulations despite their critical role in thrombotic risk. Thus, geometry represents one component of risk within a broader biologic and clinical context.

Future investigations should pursue several directions. First, expansion to a larger, prospective cohort will enable statistical validation of geometric predictors and facilitate the development of quantitative hemodynamic thresholds associated with thrombosis risk. Second, integration of computational modeling with patient-specific platelet function assays or circulating biomarkers may directly link mechanical stress exposure to biologic activation signatures. Third, incorporation of compliant wall models and advanced flow modeling techniques will enhance physiologic realism.

Ultimately, these efforts may support the development of patient-specific surgical planning tools in which preoperative imaging is used to simulate alternative graft configurations and select geometries that minimize adverse shear exposure while preserving adequate pulmonary blood flow. Such an approach would represent a shift from reactive management of thrombosis to proactive biomechanical risk mitigation at the time of shunt implantation.

## CONCLUSIONS

In this patient-specific computational study, surgically modifiable geometric features of the mBTTS shunt emerged as key determinants of thrombosis-related hemodynamics in anatomically realistic configurations. Elevated wall shear and elongational strain consistently localized to the shunt–subclavian junction across anatomically diverse patients, establishing this region as a reproducible biomechanical vulnerability. Deviation of the shunt insertion angle from perpendicular correlated significantly with local shear exposure, and larger shunt diameters were associated with concordant reductions in shear metrics, identifying insertion angle and shunt diameter as primary surgically modifiable targets. Thrombosed shunts exhibited higher cycle-averaged shear exposure than patent shunts, supporting a mechanistic link between geometric design and thrombosis risk that warrants confirmation in larger prospective cohorts.

These findings extend principles identified in idealized computational models into patient-specific anatomies under physiologic boundary conditions, providing translational validation that geometry-informed optimization may mitigate thrombogenic flow at its source. Because systemic anticoagulation cannot modify the local mechanical environment that initiates thrombosis at the shunt interface, geometric and pharmacological strategies may serve as complementary strategies. Incorporation of preoperative computational hemodynamic assessment into precision surgical planning may have the potential to reduce thrombosis risk in this vulnerable population and represents a concrete step toward precision congenital heart palliation.

## Data Availability

All data produced in the present study are available upon reasonable request to the authors

## Supplemental Material

## Abbreviations and Acronyms

CHD: Congenital Heart Disease
mBTTS: modified Blalock-Taussig-Thomas shunt
SIPA: shear induced platelet activation & aggregation
VWF: von Willebrand Factor
CT: computer tomography
MR: magnetic resonance
HLHS: hypoplastic left heart syndrome
PA: pulmonary artery
STS: society of thoracic surgeons
CFD: computational fluid dynamics
WSR: wall shear rate
ESR: elongational strain rate
Re: Reynolds number

